# A Deterministic Agent-based Model with Antibody Dynamics Information in COVID-19 Epidemic Simulation

**DOI:** 10.1101/2022.05.11.22274979

**Authors:** Zhaobin Xu, Hongmei Zhang

## Abstract

Accurate prediction of the temporal and spatial characteristics of COVID-19 infection can provide favorable guidance for epidemic prevention and control. We first introduce individual antibody dynamics into an agent-based model. Antibody dynamics model can well explain the antibody fading effects through time. Based on this model, we further developed an agent-based approach which considers the dynamic behaviors of each individual antibodies. The method can effectively reflect the dynamic interaction between the antibody and the virus in each host body in the overall population. Using this method, we can accurately predict the temporal and spatial characteristics of the epidemic. It can quantitatively calculate the number and spatial distribution of infected persons with different symptoms at different times. At the same time, our model can predict the prevention and control effect of different prevention and control measures. At present, China’s dynamic zero strategies mainly include large-scale nucleic acid test, isolation of positive infected persons and their close contacts. Our model demonstrates that for a less infectious and more virulent variant, this approach can achieve good preventive effect. The effect of reducing social contacts and quarantining only positive infected persons is relatively weaker on epidemic control. This can explain why China’s targeted epidemic-control measures had an excellent performance in 2020 and 2021. However, our model also warns that for the highly infectious and less virulent variant, targeted epidemic-control measures can no longer achieve effective control of the epidemic. Therefore, we must choose to quarantine potential infected groups in a wider range (such as the quarantine of secondary close contact and tertiary close contact) or coexist with the virus. Furthermore, our model has a strong traceability ability, which can effectively conduct epidemiological investigation to unearth patient number zero based on the early epidemic distribution. In the end, our model expands the traditional approaches of epidemiological simulation and provides an alternative in epidemic modeling.

**Major findings:** First, a method was developed to integrate the characteristics of individual antibody dynamics into epidemic prediction;

Second, this model can effectively predict the spatiotemporal characteristics of patients with different symptoms (including asymptomatic patients, mild and severe patients, etc.);

Thirdly, this model proves that China’s dynamic zero strategy which include the quarantine of close contact people is more efficient than just isolating positive cases;

Fourth: This model also reflects the limitations of targeted epidemic-control strategies and warns that for the highly infectious and less virulent variant, targeted epidemic-control measures can no longer achieve effective control of the epidemic;

Fifth, this model can help epidemiological research and find out patient zero according to the early incidence of the epidemic.

## 1 Introduction

Since the outbreak of COVID-19, it has caused great damage to global public health security and human economic life. The characteristics of COVID-19 are also constantly changing, with enhanced transmission potential which had been proved by clinical data [1-2]. This would pose a great challenge to the public prevention strategies, especially to some countries insisting the dynamic zero policy. For example, at the end of 2021, the outbreak caused by the Delta variant strain in Xi’an, China, and the outbreak caused by the Omicron strain in Tianjin seriously affected the social and economic activities within a few weeks to a month [3]. As the virus infection potential increases, more and more local outbreaks will recur in different parts of China. It is of urgent need to re-estimate the effect of targeted epidemic-control measures and the accessibility of “dynamic zero” goal.

Mathematical models can be well used to predict and assess the trend of the epidemic. At present, there are mainly two kinds of models for predicting the epidemic situation of COVID-19. One is the widely used ordinary differential equations model, which can be divided into SIR (susceptible population, infected population, recovered population), SEIR (susceptible, exposed, infected, recovered), SEIQR (susceptible, exposed, infected, quarantined, recovered), etc., according to the specific selection of the compartments. These models have good data fitting function and certain prediction ability. As early as 2017, Xiao Dongmei and others expanded the SIR model, adding the asymptomatic patient module and seasonal factors [4]. Since the outbreak of COVID-19, more and more researchers have participated in the research and improvement of the traditional model, and several typical research works include Lai Yingcheng, SHIJR (S: susceptible, H: hidden, I: infected, J: infected and confirmed, R: recovered) model, which adds a time delay function to the traditional model to more accurately predict the development of the epidemic [5-6]. Wang Hao, Jiang Jifa and others developed the SIR model of stochastic discrete time [7]. Jin Zhen et al. used the extended SEIQR model to predict the impact of Hubei imported cases in the early stage of the epidemic on the development of the epidemic in Shanxi [8], they applied this model to further analyze the impact of public policies on the development of the epidemic [9]. These studies apply and expand the traditional epidemic spread model, and enhance the applicability of the model. However, the compartment model based on ordinary differential equations also has some drawbacks, the most important of which is that it cannot take into account the heterogeneity within the system [10]. Most of these ODE (ordinary differential equation) models can achieve good fitting results. However, fitting based on the data in the early stage of the epidemic often leads to a low number of initial susceptible groups S (susceptible). At the same time, because most ODE models cannot take into account the geographical distribution characteristics and specific contact characteristics of people, they cannot accurately predict the epidemic development at a geometric level.

The second is the agent-based model, which benefits from the rapid development of computing speed. Based on the characteristics of individuals, this model can study the incidence probability of a single individual, thus summarizing the epidemic characteristics of the total population [11-12]. Compared with the traditional ordinary differential model, the individual-based model can better reflect the heterogeneity within the system. Agent-based approaches have been widely used in the fields of supply chain optimization [13], ancient civilization waning [14], and dynamic modeling of the immune system [15]. After the outbreak of COVID-19, many scholars applied it to the prediction and prevention and control of COVID-19. For example, Hoertel et al. proposed an agent-based model based on random perturbation to simulate the epidemic of COVID-19 in France in the early stage [16]. Hinch et al. established an agent-based framework called “OpenABM-Covid19” to study non-pharmaceutical interventions for COVID-19 in the UK [17]. Erik Cuevas proposed a model based on the movement of individual locations to assess the risk of transmission of COVID-19 [18]. We developed a continuous Markov chain model based on the individual model to better simulate the spatio-temporal characteristics of the epidemic development [19]. The model we developed has three main characteristics:

First, individual contact information can be fused into the model to realize epidemic prediction under the guidance of big data;

Second, it can reflect the infection probability of a single individual at different times;

Third, our model takes into account the age of individuals and the decline of immunity over time after natural infection or vaccination, and can predict the resurgence of the epidemic wave.

According to the updated characteristics of the epidemic situation, we further improved our model and endowed it with new functions. These improvements mainly include two parts:

The first part is to give the model the ability to trace the source of the epidemic and the potential transmission chain according to the current incidence of the disease. Second, to improve the accuracy of model prediction, we use our previous research results [19-20] to integrate the information of individual antibodies and viruses into the overall population network. The integration of antibody dynamics in each individual breaks the limitation in that we can explicitly model the waning effect of antibody and simulate the infection possibility based on antibody-virus interaction in a deterministic way. This will accurately predict and evaluate the risk of disease and transmission of a single individual, together with the population outcome and behaviors.

Since the late spring of 2020, on the basis of its own experience, China’s epidemic prevention and control has put forward the overall policy as “targeted control, dynamic zero” [21]. The core of this policy is to isolate the positive patients and their close contacts on the basis of nucleic acid test within susceptible infection groups. This strategy had an outstanding performance in balancing the costs and prevention effects during 2020 and 2021. However, this “targeted control” strategy gradually loses its effects in the epidemic prevention, especially toward the invasion of omicron variants. Since march 1^st^, 2022, the COVID-19 epidemic displayed a trend of rapid spread in China mainland. More and more Chinese local governments have abandoned the policy of precise prevention and control in order to completely block the infection. The purpose of this study is to establish an accurate mathematical model to predict the development and spread trend of the local epidemic. On the basis of this model, we focus on analyzing the effectiveness of different epidemic prevention policies, including “lockdown”, “quarantine of positive cases”, and “precise prevention and control, dynamic zero” which is currently implemented in China. We tried to elucidate the effects of different prevention strategies and explained why the isolation of close contact policy would lose its efficiency in the dynamic zero attempts.

## 2 Results

### 2.1 Parameter estimation based on clinical data

In the previous research, we presented a theoretical overview of antibody dynamics and proposed a method for calculating the protection period of antibodies based on this model. According to the data of clinical experiments, we can further fit the specific parameters to obtain the dynamic characteristics of antibodies within certain population. An important feature of our model is that its parameters do not involve units. This is also puzzling to many readers. The reason we don’t integrate units into those parameters is that the unit itself is not optional in the simulation. What is important is the interaction dynamics between the antibody and the virus. Furthermore, it is not feasible to acquire the units directly through experimental data. For instance, although the CT value in the nucleic acid test can reflect the amount of the virus, it is difficult to quantitatively derive virus loading amount in host body based on the detected CT value. The interaction of antibody and virus is not a simple one-to-one binding pattern. At the same time, it is difficult to quantitatively determine the absolute value of antibody concentration at different times since most experimental data are expressed by antibody titer. The core content of this model is to accurately simulate the trend of mutual change between antibodies and viruses, so it is not required to add units to the mathematical model. By adjusting different parameters, especially the mean and variance of the parameters, we can get the characteristics of antibody dynamics behavior of a population. By comparing with the known statistical data, we can estimate the distribution of antibody dynamics parameters of the whole population. Although these parameters have no direct physical meaning, it can more accurately reflect the decline of immunity in different individuals and the possibility of reinfection at different times.

We treat that the concentration of the environmental antigenic substance as a constant value. The main alternation is the difference in binding capacity of the antibody in different individual which is defined as ***k***_**2**_ in our model. The binding kinetic between the environmental antigenic substance and the antibody, which is defined as ***k***_**7**_ in our model, is also varied among different people. The antibody kinetic performance characteristics of the population obtained by using the above parameter combination are shown in Figure 1A. The simulation result has a good match with clinical reports [22]. The main factor determining the peak antibody concentration is ***k***_**2**_, while the driving force determining the antibody waning speed comes from ***k***_**7**_. The change in the overall antibody of the population over time is shown in fig. 1A, and the change in the overall protective efficacy is shown as fig. 1b. According to this model, we can judge that in the absence of virus mutation, the overall protective efficacy of an initial 100% efficacy COVID-19 vaccine in a human population (10000 human simulation) would drop to 97.21% after 100 days, 65.44% after 150 days, 39.28% after 200 days, and 28% after 240 days. This simulation results in Figure 1b are roughly consistent with the clinical data of vaccine effectiveness [23-25]. Therefore, in subsequent simulation, we use those parameters combination to reveal the epidemic development under different prevention and control measures.

**Figure 1A:**
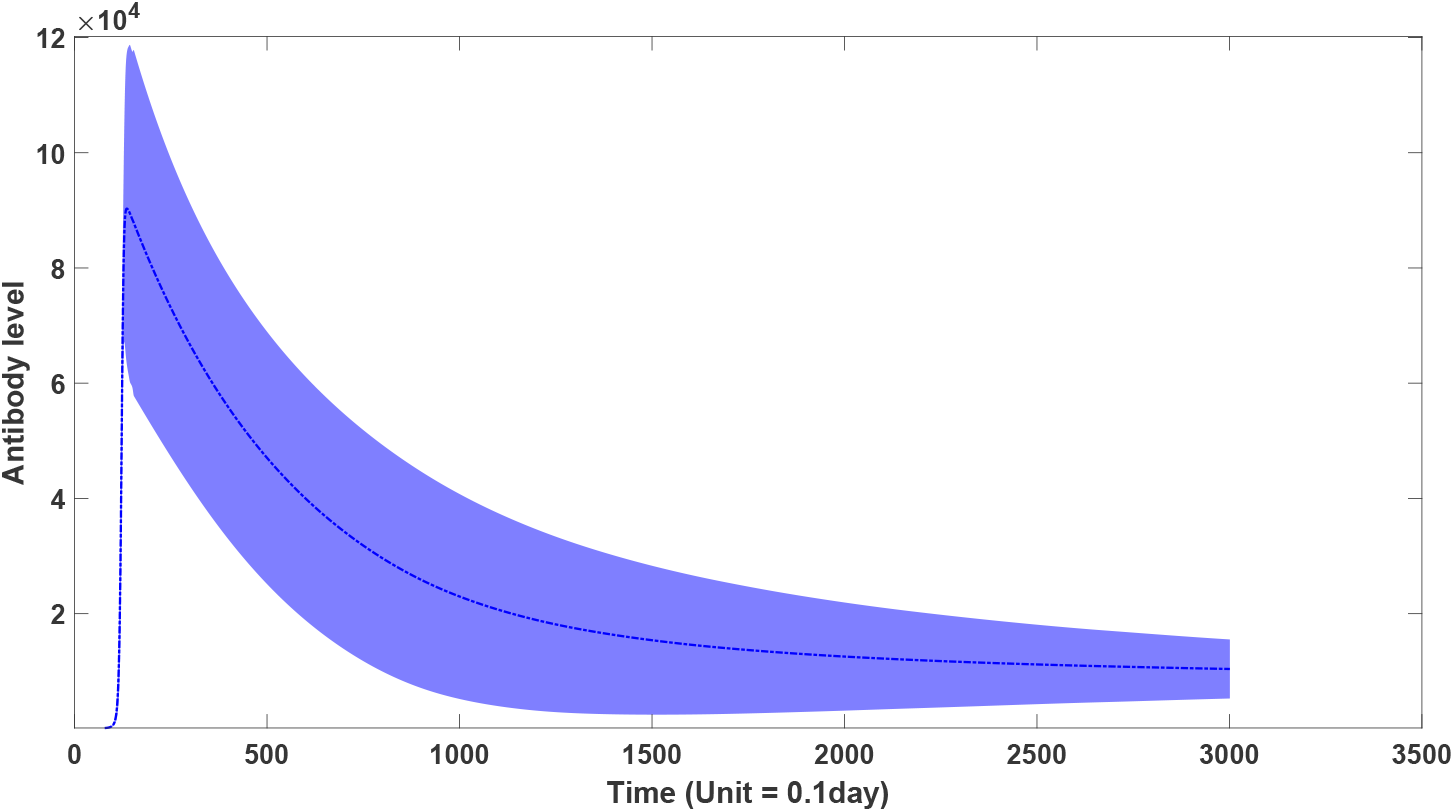
the dynamic behaviors of antibodies in the overall population through time

**Figure 1B:**
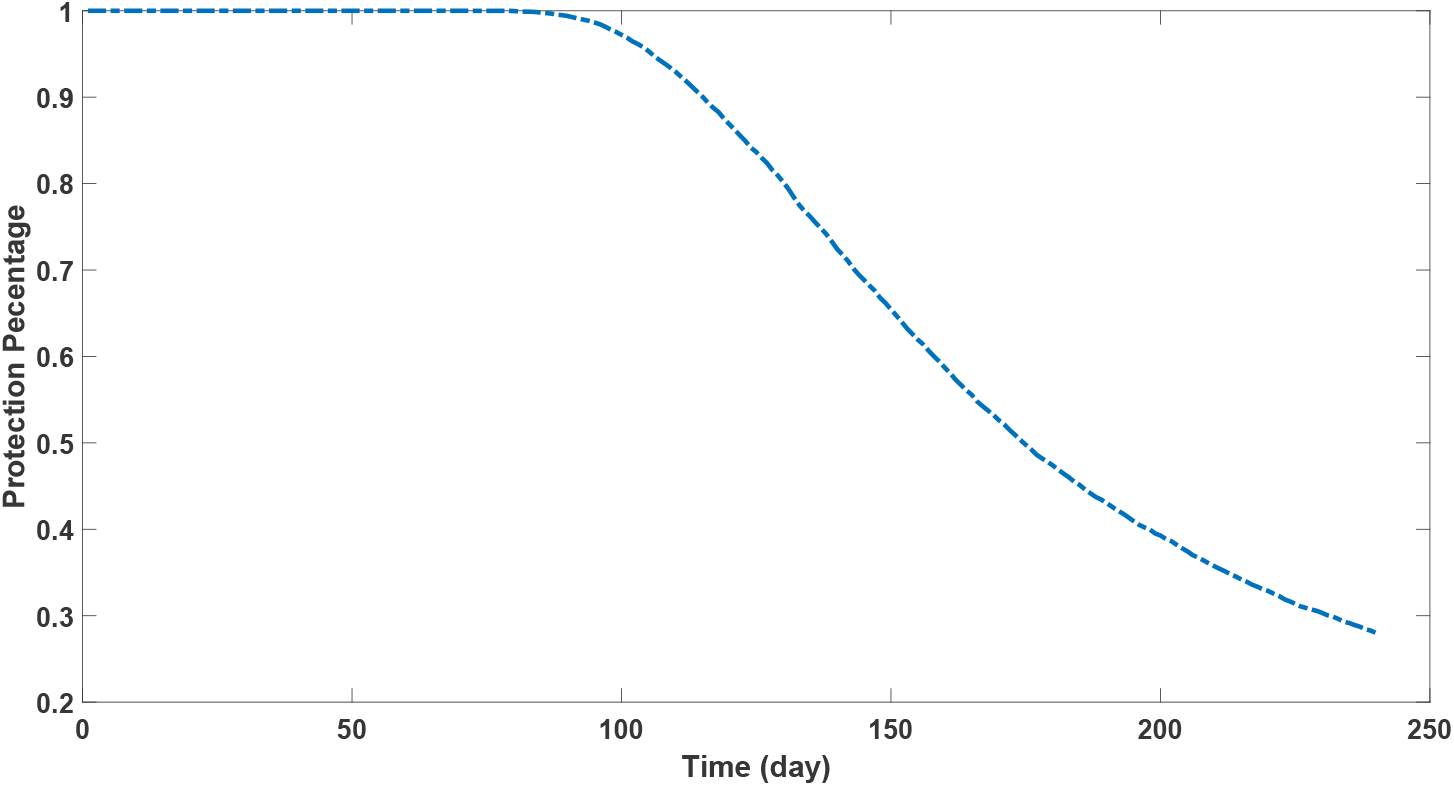
The protection performance of antibodies in the overall population through time

### 2.2 Simulation of epidemic development in a population level

Based on the parameter combination proposed in 2.1, we set up a group of 1000 people, who are randomly distributed in four regions. The contact frequency between each person and the distance between them show a certain functional relationship, which is explained in detail in our previous article [19]. This is also defined in equation (11) in the method section. A population contact matrix is derived based on people’s location. We assume that only one person who are patient zero is infected at the initial time. According to the equations (13-18) in the method part, two scenarios are simulated, one is the spread of the epidemic in the ideal state. The ideal condition assumes that the virus does not mutate through time. This ideal scenario is represented in Figure 2A. The temporal and spatial information of all infected persons is represented by Video 1A. The other one is closer to the actual epidemic development which consider the virus mutation effect. It has been demonstrated experimentally and statistically that sars-cov-2 is evolving to be milder in virulence and more feasible to be infected [26-28]. This scenario is represented in figure 2B and the temporal and spatial distribution of infections is represented in Video 1B. In Video 1, the asymptomatic infections are represented by light color with a gray level of 0.25, and the mild symptom infections are represented by medium gray level of 0.5. Severe cases are represented by dark gray with darkness scale of 1.

**Figure 2A:**
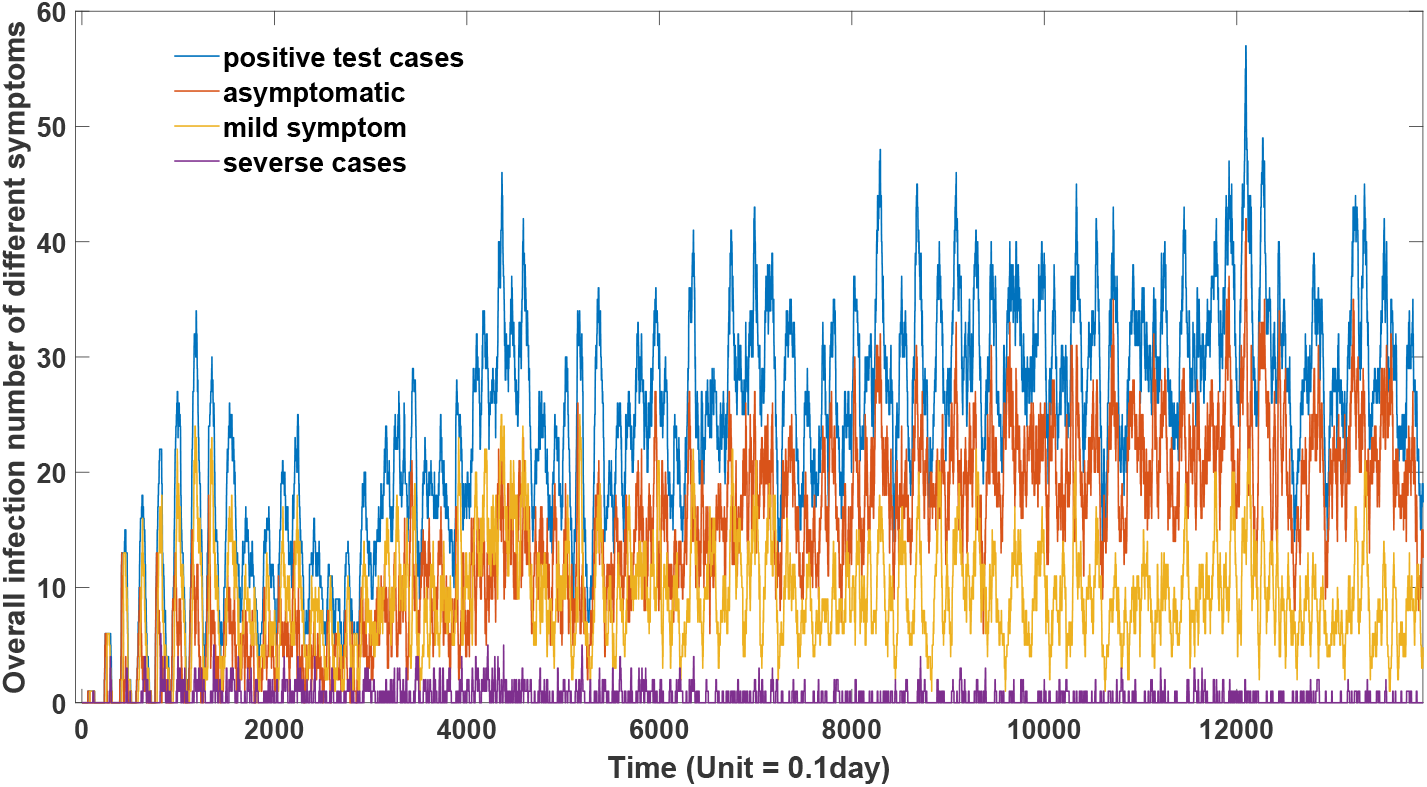
The development of epidemic in ideal condition (n = 1000)

**Figure 2B:**
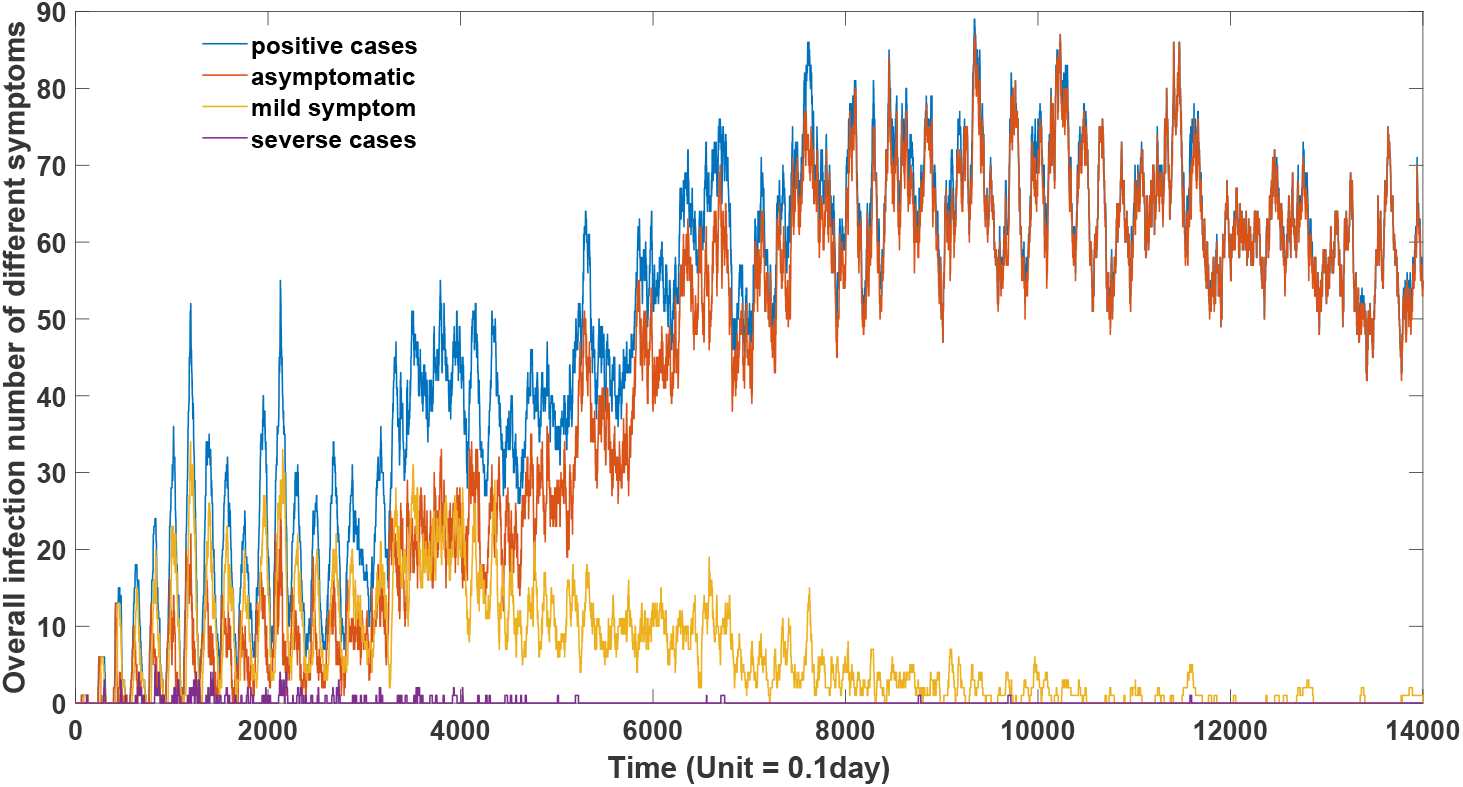
The development of epidemic in actual case (n = 1000)

This method is not only able to determine the onset time and hotspots of infection, but also quantitatively describe the transmission risk and severity of infection symptoms of each infected individual. In the model, we associate infection symptoms with virus loading in host body. In our previous research, we proposed that high viral load induces high levels of antibodies, which leads to a stronger immune response. It then shows more significant symptoms. To the contrary, low viral load could induce low levels of antibodies, resulting in a relatively mild immune response, which would display as mild or asymptomatic infection. We rationally defined several thresholds for symptoms. We defined a low threshold for a positive nucleic acid test (0.1e5 in our model), a medium threshold for a mild symptom (0.5e5 in our model), and a high threshold for a severe case (5e5 in our model). If the amount of virus in the patient is between 0.1e5 and 0.5e5, he will be defined as asymptomatic patient. Experiments have confirmed that there is a significant positive correlation between symptoms and viral content in vivo, which is reflected in the low CT value of viral amplification in severe patients [29-30]. Although there are other complex factors affecting human immune response, we believe that for the same virus, its concentration level is the main factor determining the intensity of immune response. The situation described in Figure 2B is closer to the actual situation. We can see that with the extension of time, the severity of infection will gradually reduce. This declination is due to two factors. The first one is the decrease of the efficiency of virus proliferation, that is, the decrease of virus virulence, which is manifested in the decline of k1 in our model. The second factor is due to the increased level of antibodies in the population. Because later infections are essentially reinfections (infections occurring after vaccination are also defined as reinfections), the peak load of the virus in infected persons will be relatively low, and a higher proportion of asymptomatic infected persons will appear due to the presence of increased antibodies level.

### 2.3 Evaluation of the efficiency of different epidemic prevention strategies

Based on this model, we evaluated the effect of different epidemic prevention strategies. We use a model close to the real situation to simulate epidemic situation under different prevention strategies. As described in section 2.2, this model provides a flexible virus proliferation parameter k1 with its value decline with time. Meanwhile, as indicated by clinical data, the formation of virus particles will be strongly enhanced, the transmission parameter in our model also changes over time. We evaluated the effectiveness of three common epidemic prevention methods. The first epidemic prevention policy is to increase social distance and reduce social contact frequency. This strategy can be realized by the wild application of face mask and the lockdown policy. The second epidemic prevention policy is to carry out large-scale nucleic acid tests, but only isolate the positive patients.

The third epidemic prevention policy is to carry out nucleic acid test for all people in risk areas, and isolate both positive patients and their close contacts. This strategy is described as “targeted control” by Chinese government.

We simulated the onset of disease in a small population of 1000 people over a period of 1400 days without vaccination which means that the increase in antibody was only caused by natural infection. On the first day, the outbreak was caused by the infection of patient 0. In the first six weeks, the number of infected people was small. No prevention measures were taken, and different epidemic prevention measures were taken from the seventh week.

The first epidemic prevention policy is to increase social distance and reduce social frequency. In our model, the reduction of social frequency can be accomplished by the reduction of the overall contact matrix, here we use 20% of its original value. A smaller value could be used if a more radical lock-down policy is applied. However, we also need to admit there would be more side effects under strict lock-down strategy. The performance of this policy is displayed in Figure 3. A dynamic geographic epidemic distribution is shown as Video 2.

**Figure 3:**
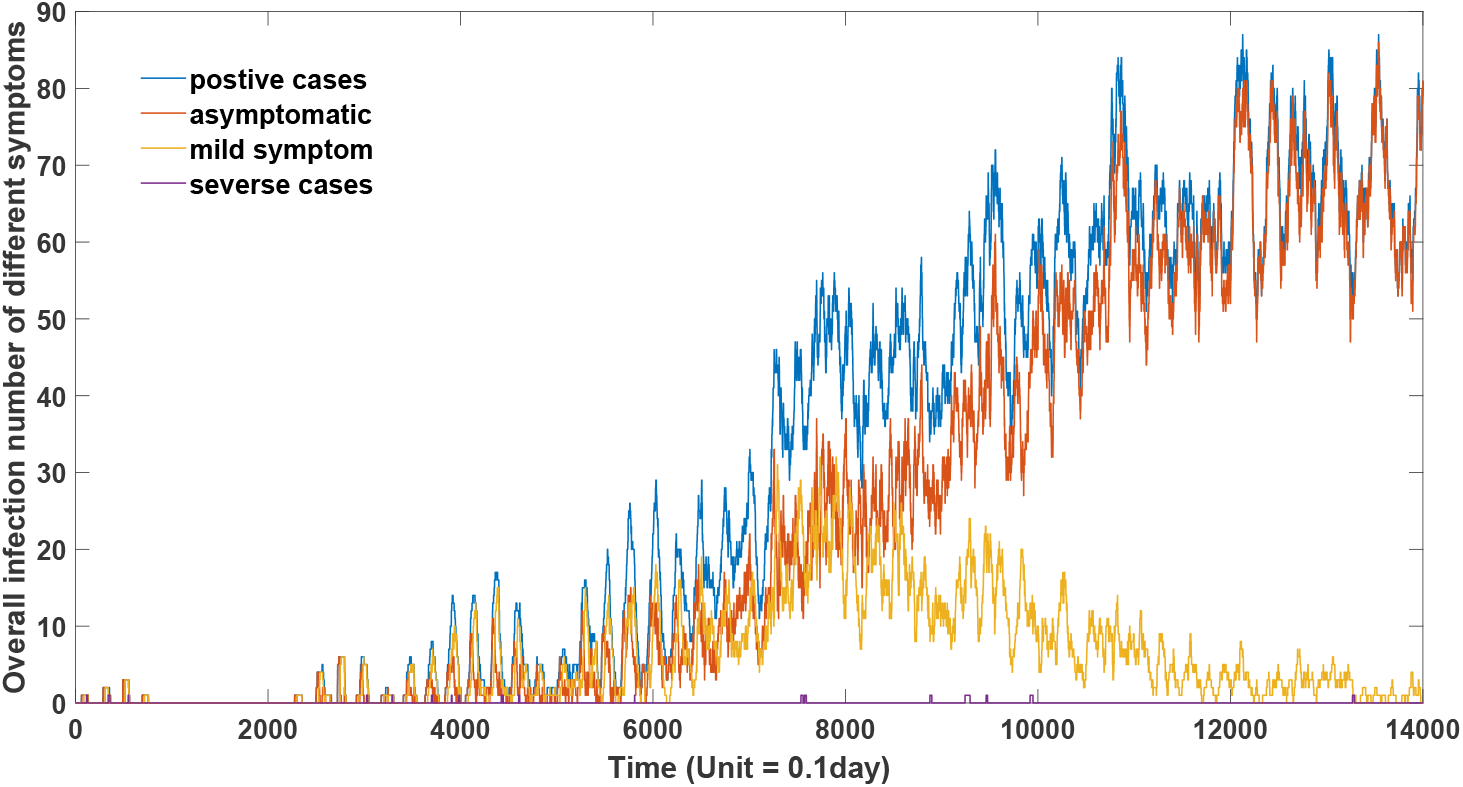
the effect of 80% lock-down on the epidemic development

The second epidemic prevention policy is to carry out large-scale nucleic acid tests, but only isolate the positive patients. Our model mimics this strategy by adjusting the contact frequency to zero for 14 days when the virus concentration in host is above 0.1e5. After 2 weeks, the original contact matrix will be restored if the virus concentration in host is below 0.1e5 which represents negative nucleic test result, otherwise an additional 14 days will be added. During the whole process, the virus loading amount inside each person in the population needs to be tracked and assessed, which means nucleic acid tests should be carried out frequently. The effect of prevention strategy is shown in Figure 4. A dynamic geographic epidemic distribution is shown as Video 3.

**Figure 4:**
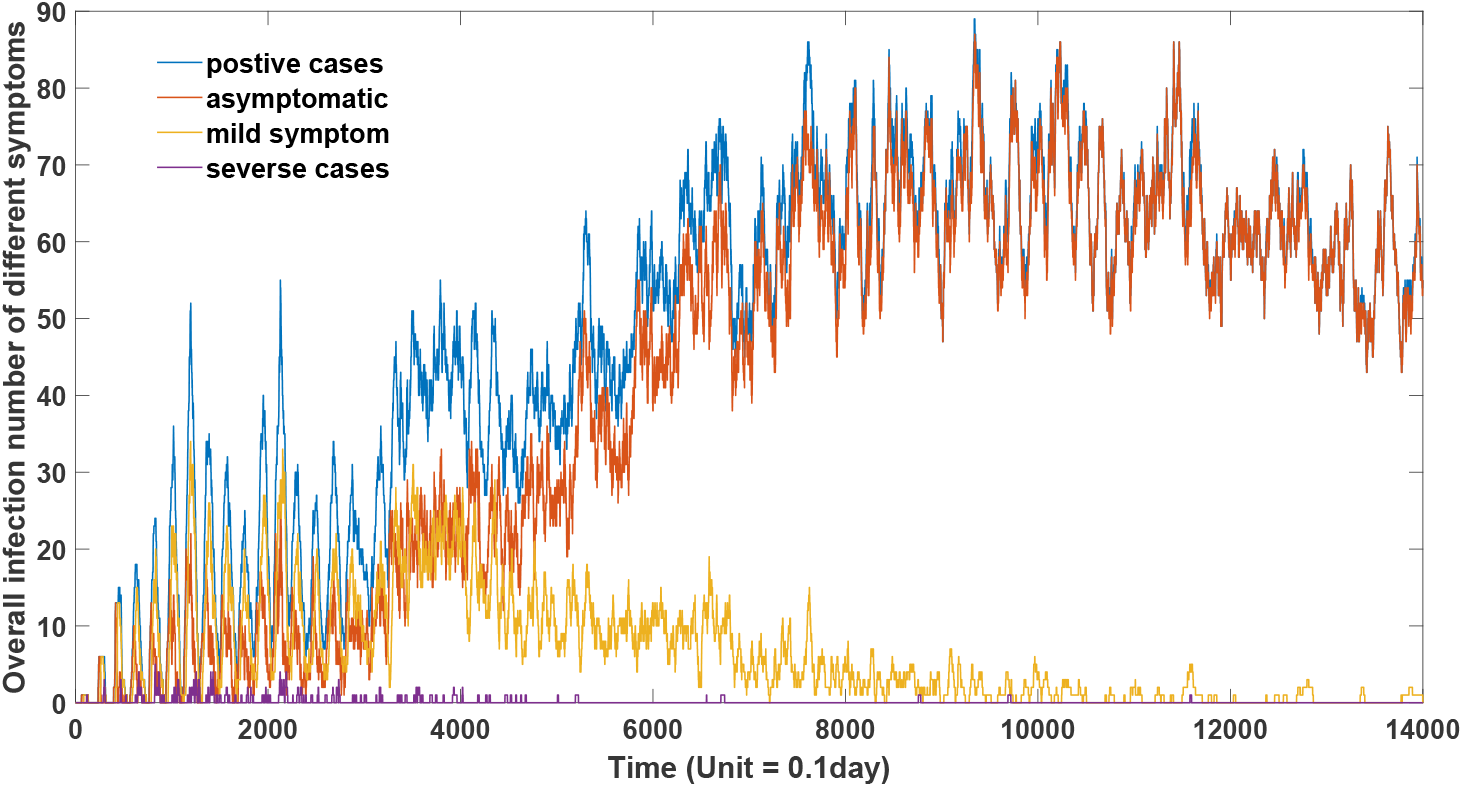
the effect of positive case quarantine on the epidemic development

The third epidemic prevention policy is to carry out nucleic acid test for all people in risk areas, and isolate positive patients and their close contacts. This strategy is defined in our model as adjusting the contact matrix of all people whose viral level is greater than 0.1 e5 to zero. Meanwhile, it also needs to change the contact matrix of close contacts to zero for 14 days. During this process, the virus level of each individual needs to be tracked and assessed which means nucleic acid tests should be performed frequently in the risk area. The effect of this strategy is shown in Figure 5. A dynamic geographic epidemic distribution is shown as Video 4.

**Figure 5:**
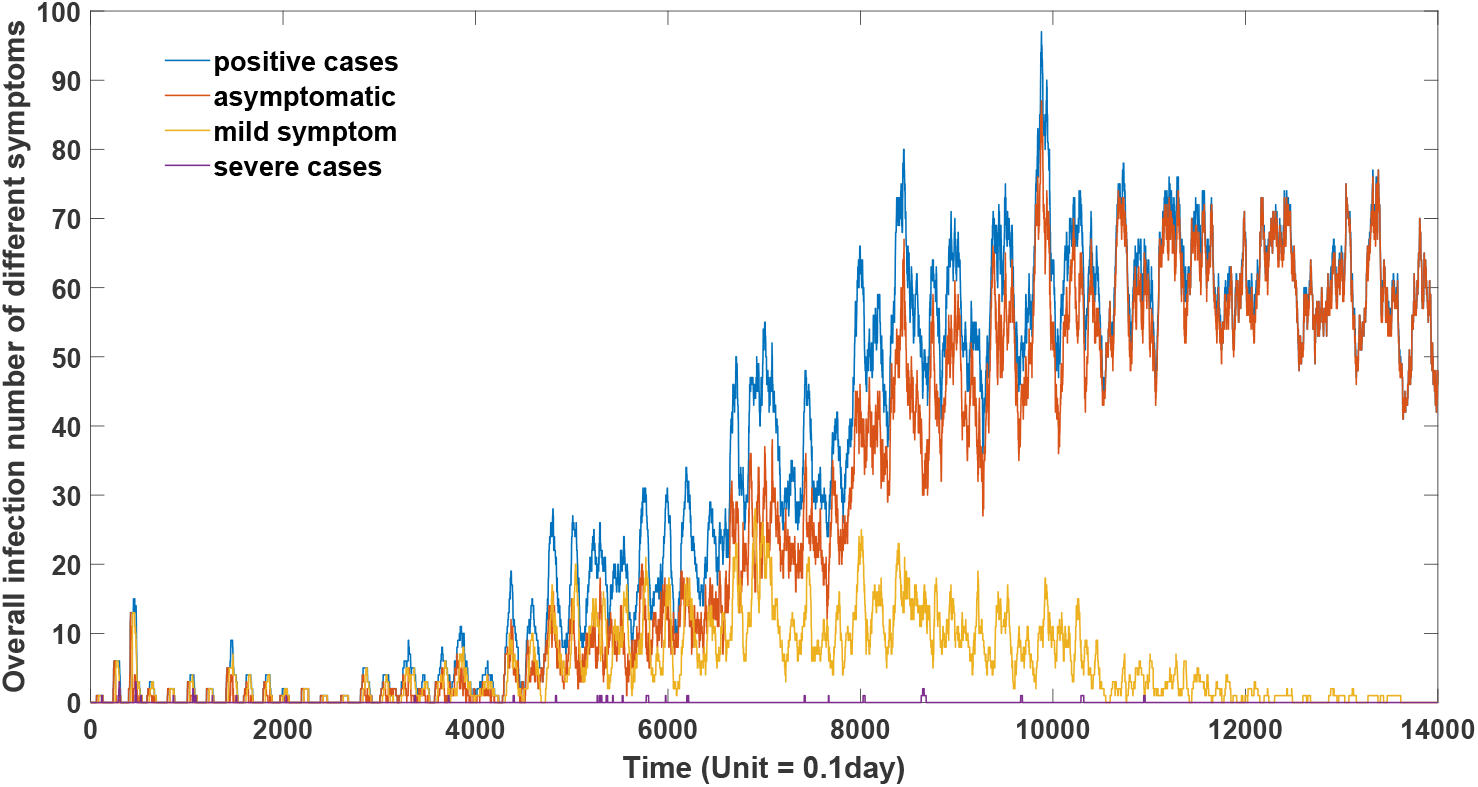
the effect of targeted-control policy on the epidemic development

For these three epidemic prevention strategies, the first one costs a lot, and the exact outcomes of this strategy is related to the strictness of the lock-down policy. Reducing social distance and contact frequency in a large magnitude can effectively control the development of the epidemic. In figure3, it can be clearly seen that when the lock-down policy reduced the contact frequency by 80%, the epidemic can be effectively controlled in the first 300 days. However, as the virus transmission capacity increases, this prevention strategy will gradually lose its effect. A more stringent lockdown is needed in order to prevent the massive epidemic outbreak. Theoretically speaking, a complete lockdown with the quarantine of overall population, can realize a complete elimination of any mutant strains. However, this approach is obviously unrealistic to be implemented. Meanwhile, we also need to consider the risk of imported cases in real scenario. Figure 3 also shows that the effects of lockdown are different for mutants with different transmission abilities. For strains with high transmission ability, although the prevention and control measures such as lockdown and wearing masks may postpone the large-scale outbreak of the epidemic, they can neither change the trend of this outbreak nor reduce the peak infection number. Given the cost of the lock-down, we do not believe that this is a long-term feasible solution. The second strategy is to carry out nucleic acid test for all the people in the outbreak area in order to screen out the positive cases. This prevention strategy only quarantines the positive patients. The second policy is much less costly and feasible to be implemented. Many countries have adopted this method in their fight against SARS-CoV-2. However, the effect of prevention and control is very poor as can be seen from Figure 4. Even for the variant with weak transmission capacity, this measure cannot efficiently inhibit the spread of the epidemic. It’s performance in the early stage is significantly worse than that of the first and third methods, especially on reducing the overall number of severe cases. The third strategy is to carry out the nucleic acid test in risk area, and isolate the positive patients and their close contacts. Compared with the first approach, the cost of the third strategy is relatively small, but the effect of prevention and control is also very considerable as can be seen from Figure 5. This targeted-control strategy can effectively control the epidemic in the first 300 days. This can explain why China’s targeted-control strategy can realized the dynamic zero goal in 2020 and 2021. However, it should be pointed out that although the isolation of close contact is better than the former two strategies, its control capacity is not unlimited but dependent on the transmission capacity of virus. The prevention effect of this strategy will gradually decline with the increase of virus transmission. With the evolution of the virus, the transmission potential of this virus will be further enhanced. The strategy of isolating close contact and positive patients will not be able to achieve the clearing effect toward those highly transmissible mutants. As can be seen from Figure 5, after 300 days, the epidemic will spread on a large scale. Therefore, the effectiveness of this policy depends on the virus transmission capacity. We should have a different expectation of this strategy on different phase of this COVID-19 epidemic. If the transmission of the virus is significantly enhanced, to ensure the effectiveness of its prevention and control, merely isolating those close contacts cannot effectively inhibit the epidemic surge. It is necessary to quarantine subjects in a broader level, such as isolation of secondary close contact and so on. This will also increase the difficulty and cost of this prevention endeavor. With the decline of virus toxicity, the practical value of prevention has gradually disappeared. People are more worried about the emergence of a more toxic strain in the future. Public health expects also concerns that the large-scale spread of the virus will lead to the emergence of highly virulent variant. This concern has dominated epidemic prevention and control policies in some countries and regions. However, our previous studies have predicted the delicate trade-off between virulence and transmission in the evolution of SARS-CoV-2. Our theory mathematically revealed the evolutionary direction of COVID-19, and theoretically proved that its future virulence would not be significantly enhanced [26]. Combined with our deterministic agent-based model, the concept of precise prevention and control for highly infectious and low-virulent viruses has been demonstrated to be unable to effectively control the virus. It is also unrealistic to adopt a large-scale quarantine policy for a long time. Therefore, we believe that at this moment, we should coexist with COVID-19 and gradually eliminate people’s fear toward COVID-19 infection. At the same time, we should also give credits to the early prevention and control measures, because effective prevention and control measures can reduce the impact of highly toxic variant in the early epidemic phage. Both the first strategy and third strategy can greatly reduce the proportion of severe cases and the overall number of deaths in the population, which is reflected in the comparison between Figure 5 and Figure 2.

### 2.5 Epidemiological investigation to unearth patient number zero based on the early epidemic distribution

Although SARS-CoV-2 has developed into a highly infectious and less virulent virus variant, precise prevention and control strategy will never lose its value as there will be new viruses with high virulence and controllable infectivity in the future. An important part of targeted-control strategy is to excavate the origin of the epidemic and the hidden transmission chains according to the known epidemic distribution. This epidemiological investigation can help discover the potential positive cases and the close contacts based on them. Therefore, it is very important to mine patient zero according to the early epidemic information. Our model provides a systematic approach on how to excavate the first infection and the elucidation of complete transmission chain on a more scientific level.

According to the population contact matrix and the epidemic distribution in early epidemic phage, we can quantitatively determine the probability of patient zero for each individual. We use the following example to demonstrate the traceability of our model. The epidemic distribution of 1000 people is shown in Figure 6A. This distribution is the infections distribution with symptom information at 30 days (300-time units) when No.1 is firstly infected in this population. This morbidity landscape consists of three severe patients, two mild patients and one asymptomatic patient. Their exact location is shown in Figure 6A.

**Figure 6A:**
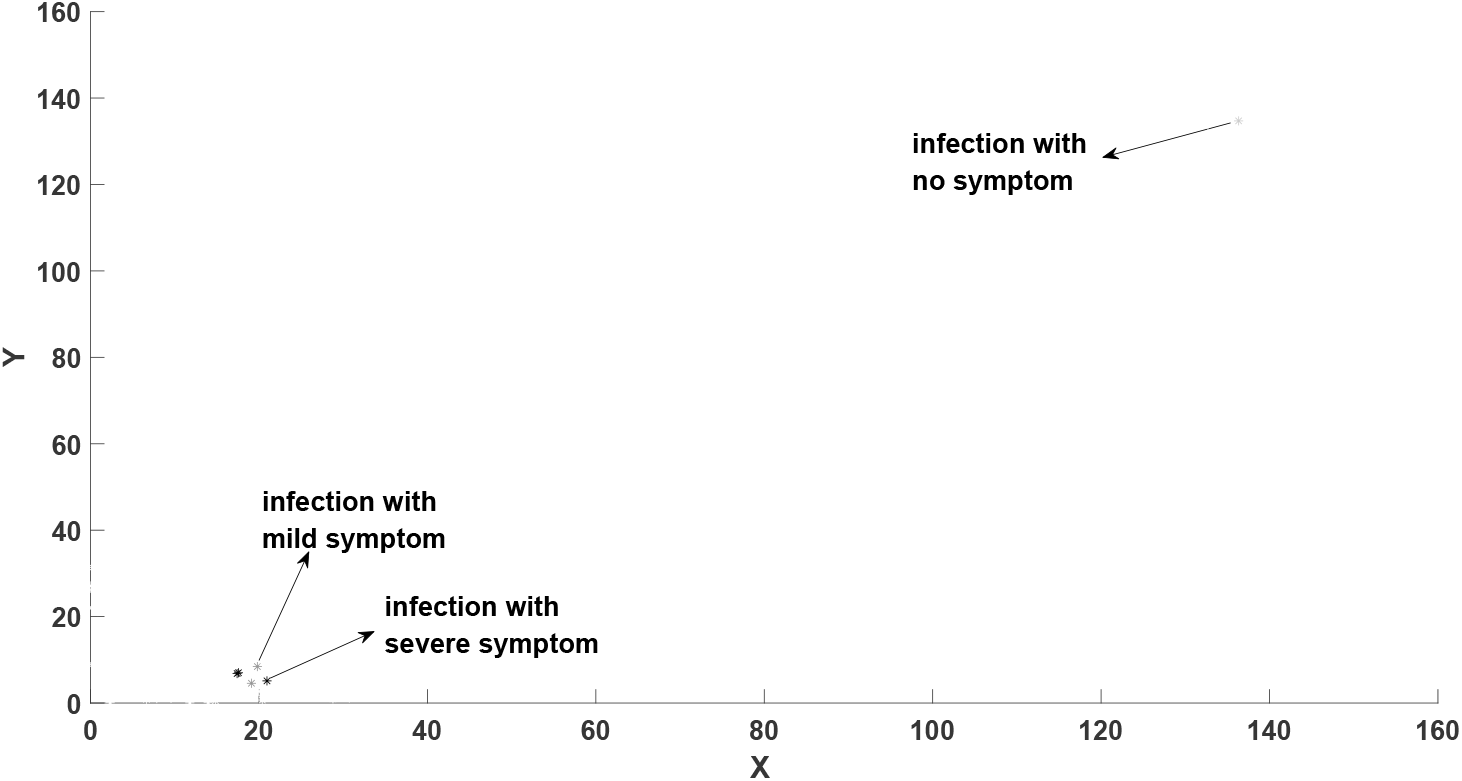
Location of patients with symptom information

We use the methods described in section 3.4 to trace the origin of this epidemic. The results are shown in Figure 6B which displays the probability of patient zero for each individual in the overall population.

**Figure 6B:**
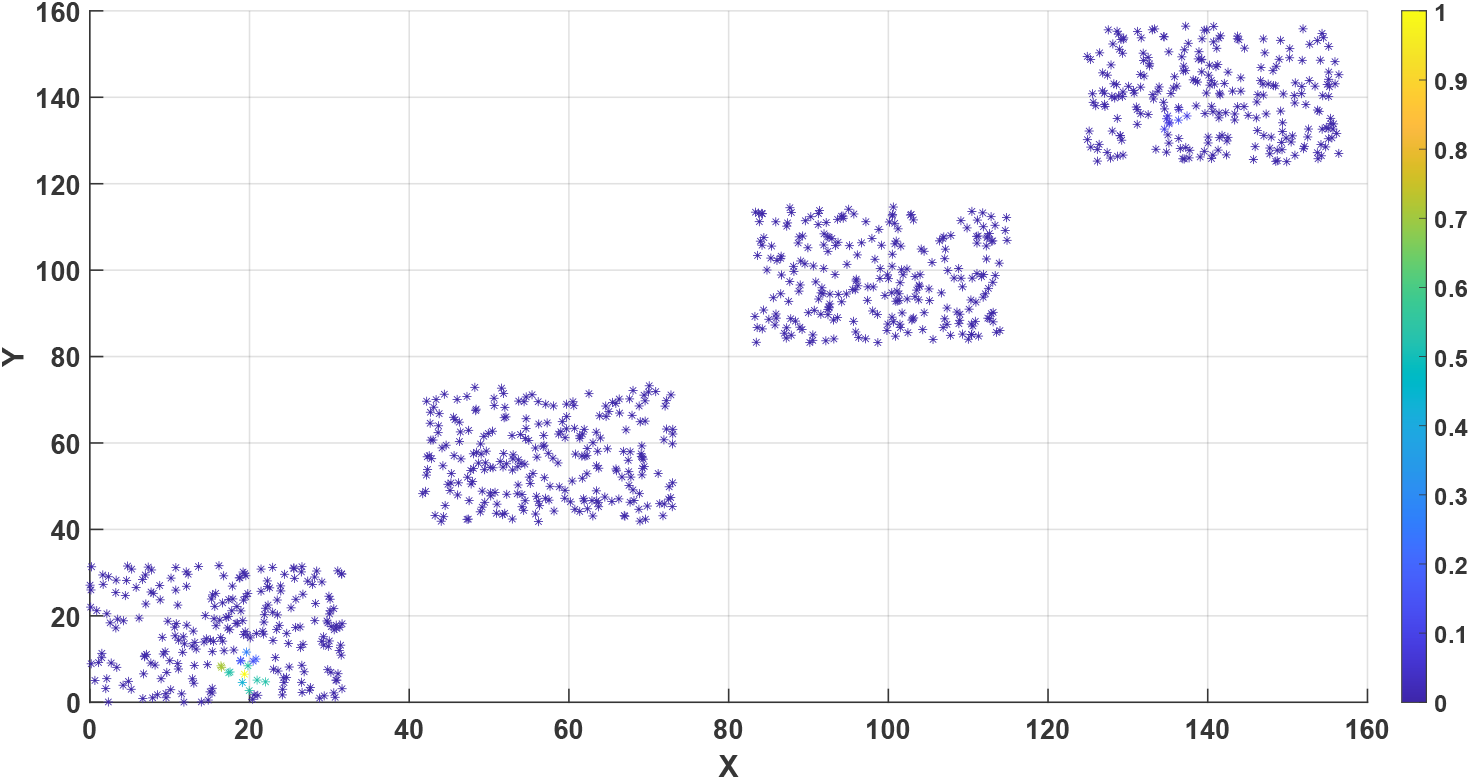
the probability of patient zero for each individual in the overall population.

This method can accurately identify patient zero and the transmission chain of the virus. As reflected in Fig. 6B, the probability that individual No.1 is the highest (100%), which is a good restoration of the real situation.

## 3 Methods

### 3.1 A review of antibody dynamic model

A simple mathematical representation of the immune response is described in the diagram below

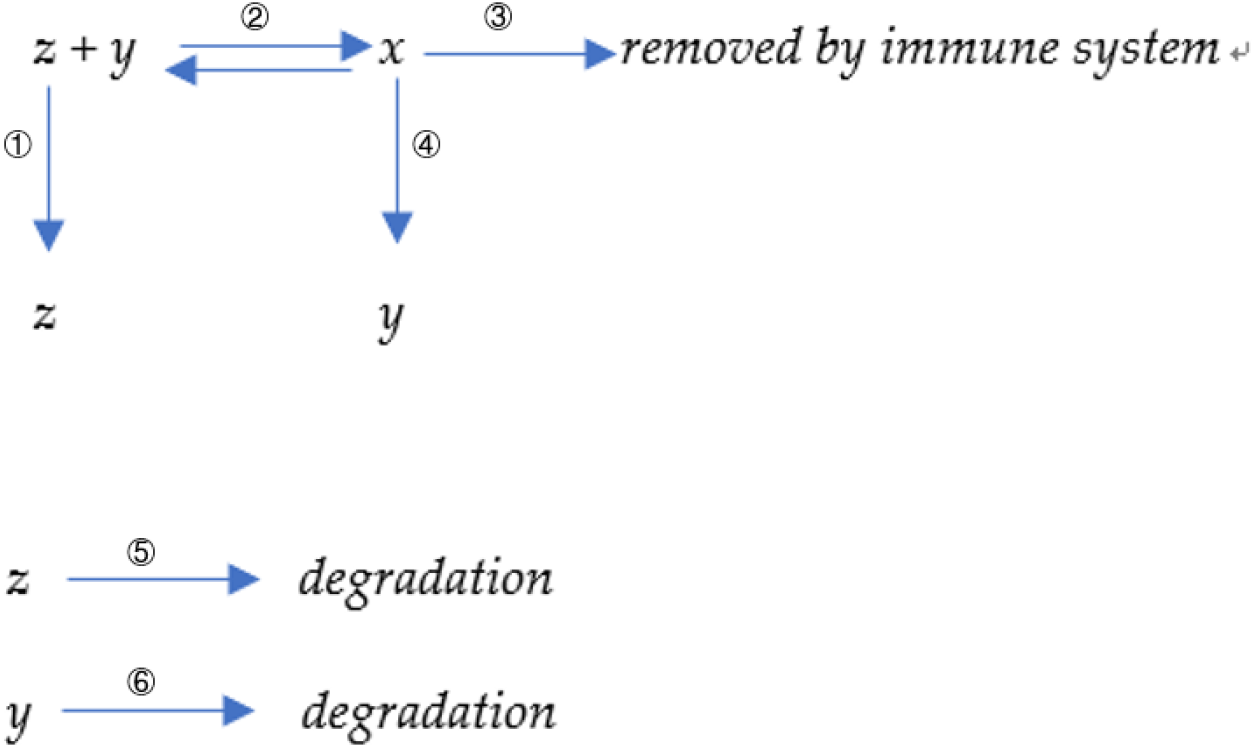

Here x denotes the amount of antibody-antigen (virus) complex, y denotes the total number of antibodies, and z denotes the number of viruses. Six processes are displayed in our model. The first reaction represents the proliferation or the replication of virus with a reaction constant ***k***_**1**_. The second reaction represents the binding reaction between virus and antibody, with a forward reaction constant ***k***_**2**_ and reverse constant ***k***_―**2**_. The third reaction represents the removal of antibody-virus complex with a reaction constant ***k***_**3**_. The forth reaction represents the induction of new antibody by the antibody-virus complex with a kinetic constant ***k***_**4**_. In immunology, those virus-antibody complexes are on the surface of B cells since the antibody are initially produced by B cells and will attach to the plasma membrane of B cells. Those complexes would further bind to the helper cells because the antibody has another structure binding region toward those receptors. Those helper cells will present the antigen part, which is a virus in this case, to the T cell. The physical placement should be B cells bind to those helper cells and further present themselves close to T cells. The T cells will handle those antigen substances; if those substances are not self-originated, they would secret signal molecules to promote the proliferation or the division of B cells who attached on them. Therefore, B cell finally get proliferated, so are the antibodies generated by their B cell. The fifth reaction represents the degradation of virus with a constant ***k***_**5**_. The sixth reaction represents the degradation of antibody with a rate constant ***k***_**6**_.

The differential equations based on those reactions are derived as below:

We established the following equations to represent the proliferation process of antibodies

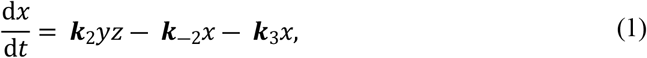

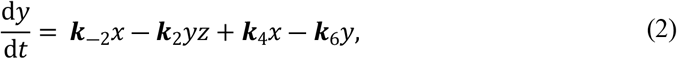

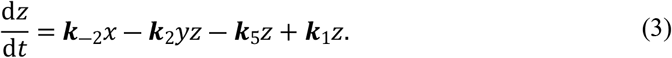

The equations(1-3) describe antibody dynamics in the presence of the associated virus. In reality, other environmental antigen can induce antibody coupling and production. To further account for this possibility, we adda new set of equations as shown below:

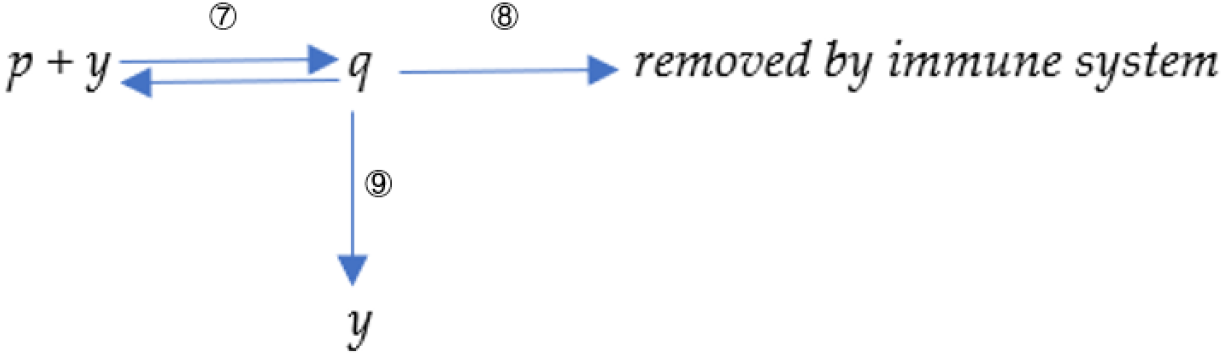

p denotes antigen-like substances in the environment; q denotes antibodies bound to antigen-like substances in the environment. Reaction 7 represents the binding reaction between antibody and environmental antigenic substances with a forward constant ***k***_7_and a reverse constant ***k***_―7_. Reaction 8 represents the remove of antibody-antigen complex q with a reaction rate ***k***_3_. Reaction 9 represents the induction of new antibody by q. Therefore, a new set of equations are derived as following:

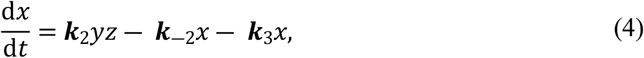

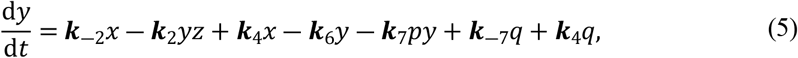

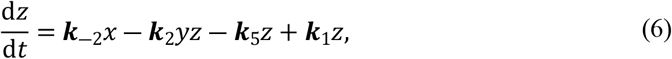

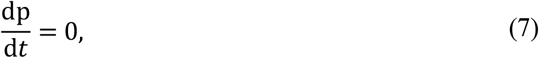

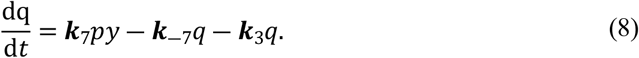

Based on the first model described in equation (1) to equation (3), the antibody level will eventually drop to zero. Since the decaying coefficient ***k***_**6**_ is bigger than zero, antibody will finally fade away in a short time. However, as we all know, some antibodies can persist in the human body for a long time and provide life-long protection. It forms the basis for a vaccine. Immunologists called the long-lasting B cell or T cell as “memory cell”. Experiments gradually demonstrated that although the so-called “memory cells” are some specific forms of immune cells [31-32], they have the similar half-lives as normal CD8+ cells [33]. Therefore, maintaining antibody level from “memory cells” should be explained as there is a stimulation that could trigger the proliferation of memory cells all the time. We suppose this stimulation derives from the existence of environmental antigen-like substances. We tend to agree that neutralizing antibody, no matter how specific it is, could have weak binding with other macromolecules in the solution. Those weak binding partners are defined as “environmental antigen-like substances”, they could be food-resourced, air-resourced, or even self-resourced. The presentation of those substances to T cell would give weak signals to proliferate the memory B cells or T cells. Because the binding is weak, and the protein sequence of environmental antigen-like substances is ordinarily close to our own body and less antigenic, the stimulation signal secreted by T cell is correspondingly weak. It would reach a balance at a certain time point when the decaying of memory cells is equal to the new generations.

### 3.2 A deterministic agent-based model with antibody dynamics information

We further integrate the antibody dynamics information into the agent-based model. The continuous Markov-chain model of epidemic simulation was proposed in our previous work [19]. It is assumed that there are N individuals in a population and there are different contact probabilities among those individuals. The infection probability is positively correlated with contact probability. For the simplified model, the relation constant is 1, which means the infection probability equals contact probability. The contact probabilities with themselves are zero. In this way, a matrix with N column × N rows is established, which has the following characteristics

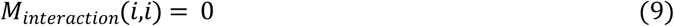

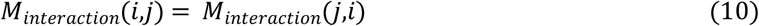

where *M*_*interaction*_(*i,j*) stands for the interaction possibility between individual *i* and individual *j*. An accurate contact matrix can be obtained by tracking the individual contact probability in a natural population group. For example, each person’s mobile phone can be recorded to obtain the population contact matrix within a particular time phase. The contact matrix is temporal and dynamic, which means it changes over time. However, it is difficult to obtain such accurate data at present. Therefore, the contact frequency is determined according to the relative distance between individuals, as shown in Equation (11).

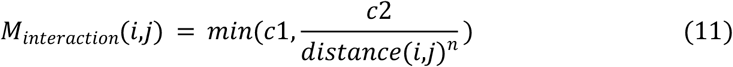

where *c1* is the maximal contact possibility between agent *i* and agent *j*. In particular, the values of *c1, c2*, and *n* are preliminarily determined according to the initial reproduction constant *R*_*0*_ of the virus. According to the contact matrix, we can further determine the number of environmental invasive viruses received by a specific individual in a specific period of time.

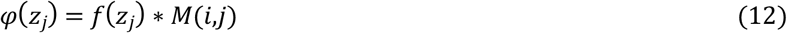

An extensive set of ordinary differential equations is further constructed. Assuming that the number of individuals in the population is *N*, the antibody virus complex in each individual is represented as *x*_*i*_, the concentration of antibodies is represented as *y*_*i*_, virus concentration is represented as *z*_*i*_, environmental antigenic substances is represented as *p*_*i*_, and the antibody-environment antigen complex is represented as *q*_*i*_.

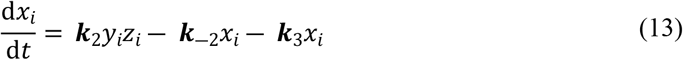

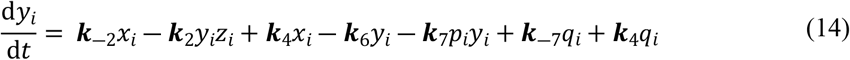

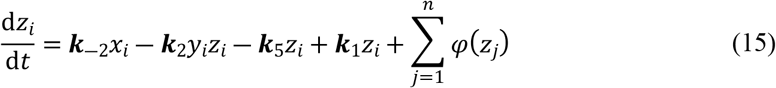

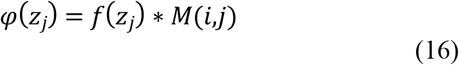

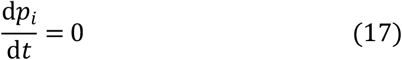

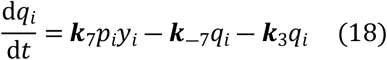

This system of equations contains *5N* variables, where the last term in equation (15) 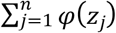 represents the overall number of viruses transmitted to this individual by other individuals in the whole population. *φ*(*z*_*j*_) is described in Equation (16). *φ*(*z*_*j*_) is equal to the number of viruses *f*(*z*_*j*_) released by individual *j* times its contact frequency *M*(*i,j*) with individual *i*. We can generate random kinetic parameters that conform to the real population distribution through the first step of simulation. For such a large system of ordinary differential equations, we can solve it quickly by increasing the step size.

For the transformation of the simulation results (converting individual antibody and virus levels into individual morbidity and total population morbidity), we set different thresholds to screen out the infections with certain symptoms and calculate the corresponding infection population of different symptoms. For the nucleic acid test, we assign a threshold to distinguish positive and negative case. When the value is greater than this threshold, individual will exhibit a positive nucleic acid test result, we define this threshold as 0.1e5. We further define asymptomatic patients range from 0.1e5 to 0.5e5; mild symptoms with range from 0.5e5 to 5e5 and severe cases range from 5e5 to∞. After correlating the virus quantity in host cell to the displayed symptom, we can get the overall epidemic development characteristics of specific groups.

### 3.3 Estimating the effects of different prevention strategies

Three prevention strategies were evaluated using this model.

1. Reduce social contact frequency In our model, we multiple the population interaction matrix with a factor *f* which ranges from 0 to 1 to mimic the lock-down effect. The contact frequency of the whole population engages a non-discriminatory declination. The stricter the policy, the smaller the *f* value.

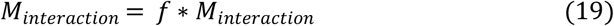
2. Extensive nucleic acid testing and quarantine positive cases The pseudocodes are presented as following: **for *i = 1:M* (*M* represents the overall steps)** **for *j = 1:N* (*N* represents the overall number of people in the population)**

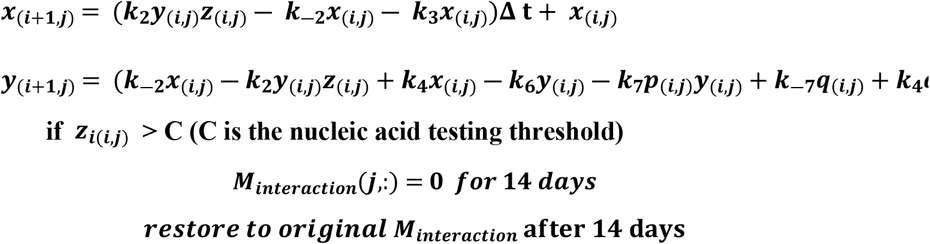

**end**

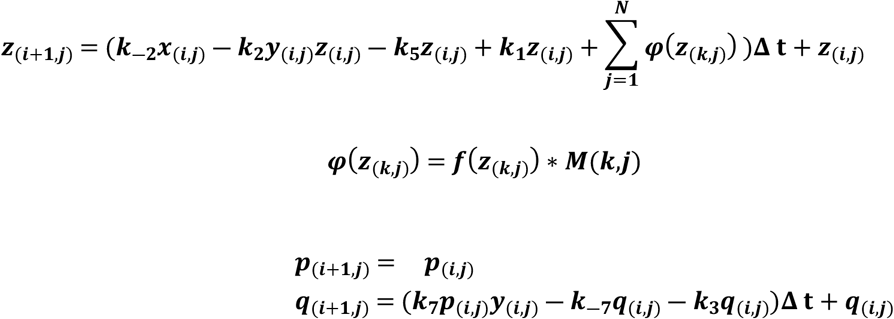

**end** **end** *x*_(*i,j*)_ represent the antibody-virus complex level of individual *j* at *i-th* time point. *y*_(*i,j*)_ represent the antibody level of individual *j* at *i-th* time point. *z*_(*i,j*)_ represent the virus level of individual *j* at *i-th* time point. *p*_(*i,j*)_ represent the environmental antigen level of individual *j* at *i-th* time point. *q*_(*i,j*)_ represent the antibody-environmental antigen complex level of individual *j* at *i-th* time point.
3. Extensive nucleic acid testing and quarantine both positive cases and their close contacts The pseudocodes are presented as following: **for *i = 1:M* (*M* represents the overall steps)** **for *j = 1:N* (*N* represents the overall number of people in the population)**

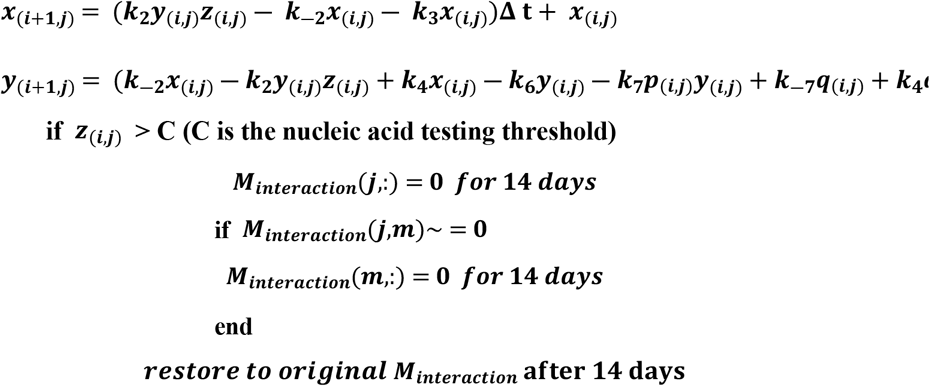

**end**

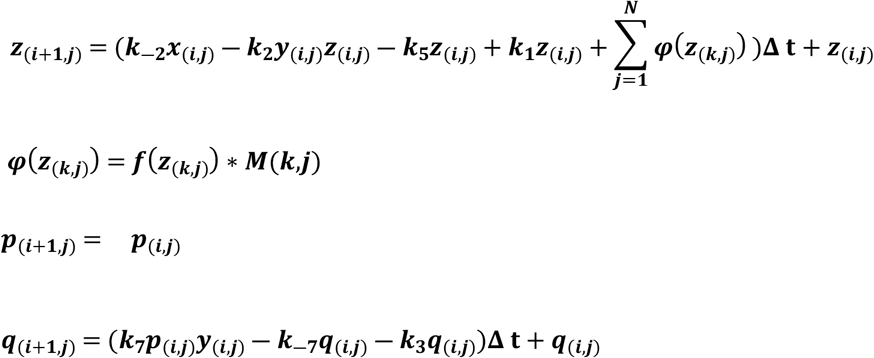

**end** **end** *x*_(*i,j*)_ represent the antibody-virus complex level of individual *j* at *i-th* time point. *y*_(*i,j*)_ represent the antibody level of individual *j* at *i-th* time point. *z*_(*i,j*)_ represent the virus level of individual *j* at *i-th* time point. *p*_(*i,j*)_ represent the environmental antigen level of individual *j* at *i-th* time point. *q*_(*i,j*)_ represent the antibody-environmental antigen complex level of individual *j* at *i-th* time point.

### 3.4 Tracing the epidemic origin based on current infection information

We can obtain a morbidity landscape of the overall population at different time point. Each individual can be assigned to different value based on their current symptom. For instance, we assign 1 to severe case, 0.5 to mild symptom, 0.25 to asymptomatic infection and 0 to negative cases. Therefore, a matrix *S* can be generated which can represent the symptoms of all individual at different time point. *S(i,j)* stands for the specific symptoms of individual i at j-th time point. The actual epidemic distribution can be represented as *S*_*target*_. We tried to find out the probability of each individual to become original zero patient. This group probability can be represented as a vector *P*.

The pseudocodes are presented as following:

**for *i = 1:N* (*N* is the population size, *i = k* stands for the individual *k* is the zero patient) for *j = 1:M* (*M* is overall step sizes or the overall time points)**

***S(i***,***j)* (It can be derived based on equation(13) to equation(18))**

***Pro(i***,***j) =* cos_similarity(*S(i***,***j)***, ***S***_***target***_)

**end**

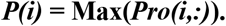

**end**

cos_similarity(*S(i,j), S*_*target*_) stands for the cosine similarity between two vectors. *P(i)* stands for the probability of individual *i* to become original zero patient.

## 4 Discussion

This study is a further study based on our previous research. A Markov chain model has been built to predict the epidemic development [19]. The Markov-chain model has several advantages over the traditional ODE models and other agent-based models. It can treat individual as an explicit object. After integrating the contact information within this population, it provides a more accurate and reliable prediction of the epidemic development. This Markov chain model theoretically denied the traditional herd immunity theory in COVID-19. It can effectively predict the multiple epidemic waves following the principle that immunity would decline through time after initial infection or vaccination. Those predictions are in good agreement with actual situations. However, a critical limitation of the Markov chain model is the antibody waning effects does not follow simple mathematical function. As shown in Figure 1B, the declination of protection effects brought by antibodies cannot be represented as simple mathematic equation. Therefore, we developed an antibody dynamic theory and attempted to integrate the antibody information into the prediction of population morbidity. Our antibody dynamics model can well explain the protection cycle of vaccines or primary infection, it can also be used to predict the protection duration of natural infection or vaccination [20]. The application of ordinary differential equations is much more accurate and physically reliable compared to the usage of simple math functions. This model is a deterministic model which could generate a fixed morbidity landscape given specific population contact matrix and the antibody dynamic parameters for each individual within certain group. The antibody concentration together with the virus loading amount within host body can more explicitly and accurately reflect the probability of infection and the number of viruses released to the environment at different time points, which provides a significant improvement toward our previous Markov chain model.

Meanwhile, this deterministic agent-based model inherits the benefits of Markov-chain model. Each individual is affected by the state of its surrounding agents by the application of contact matrix. He can spread the viruses after infection and can also receive the environmental viruses released by his contacts. Two critical features of this model can be summarized. The first character is this model can provide a more vivid morbidity landscape, including the antibody levels, the virus levels, the spatial distribution and the symptoms classification of each individual at any time point. The other character of this model is it has good tracing capacity in digging out the epidemic origin. For a small-scale population, provided the recent population contact information and morbidity status, it can help quantify the patient-zero probability for each individual. Therefore, it can further help discover the hidden transmission routines and the possible future-positive cases. It has a great potential to be used in the epidemiological investigation and epidemic control.

More than that, our model could provide some suggestions on the epidemic control strategies after we provided a quantitative evaluation of their prevention effects. Our study indicated that strict lockdown policy can effectively control the spread of the epidemic, a relaxed lockdown policy might not be able to effectively control the spread of the epidemic. The effect of lockdown is also related to the transmissibility of the virus: when the variant with enhanced transmissibility appears, previously successful lockdown measures may lose their effectiveness in epidemic control. Considering the enormous cost of large-scale lockdown, we do not recommend this strategy in future infectious disease control. Large-scale nucleic acid test and the quarantine of positive patients cannot effectively control the epidemic. However, when we expand the quarantine subject to close contacts, it could achieve a much better prevention effect. This can explain why the Chinese “targeted-control” policy achieved an excellent effect in 2020 and 2021. Lockdown and targeted-control can greatly reduce the overall proportion of severe cases and deaths in the overall epidemic. Therefore, our model confirms and appreciates the effectiveness and feasibility of China’s targeted-control policy in the prevention of highly toxic diseases, which effectively avoids the occurrence of a large number of severe and death cases. Nevertheless, we also need to realize this target-control strategy is not omnipotent, this strategy will gradually lose its effect with the increase of virus infectivity. When the transmission of the virus has been greatly enhanced, the control effect of the targeted-control strategy has become no longer excellent. Combined with our previous theory of virus virulence evolution [26], we believe that China should cancel the goal of dynamic zero at this stage. A firm pursuit of dynamic zero target will inevitably lead to large-scale lockdown instead of “targeted-control”. This large-scale lockdown is not sustainable.

## Data Availability

All data produced in the present work are contained in the manuscript

## Supplementary Materials

Videos 1to4 are displayed in supplementary materials. Matlab codes can be accessed at: https://github.com/zhaobinxu23/antibody_dynamics_agent-based_model

## Author Contributions

Conceptualization, Z.X.; methodology, Z.X.; writing—original draft preparation, Z.X.; writing— review and editing, H.Z. and Z.H.; funding acquisition, Z.X. All authors have read and agreed to the published version of the manuscript.

## Acknowledgments

We thank Dr. Dongqing Wei from Shanghai Jiao Tong University for fruitful initial discussions. Yushan Zhu from Tsinghua University, Qiangcheng Zeng, Zhenlin Wei and Tianxiang Chen from Dezhou University for helpful conversations, comments, and clarifications.

